# Linezolid population pharmacokinetics in rifampicin-resistant tuberculosis patients and interpretation challenges of the probability of target attainment

**DOI:** 10.64898/2025.12.04.25340900

**Authors:** Bern-Thomas Nyang’wa, Ilaria Motta, Ronelle Moodliar, Varvara Solodovnikova, Shakira Rajaram, Mohammed Rasool, Catherine Berry, Geraint Davies, David Moore, Frank Kloprogge

**Affiliations:** Médecins sans Frontieres, Public Health Department, Amsterdam, The Netherlands; University College London, MRC-CTU, London, United Kingdom; THINK (TB & HIV Investigative Network) Durban, South Africa; Republican Scientific and Practical Centre of Pulmonology and Tuberculosis, Minsk, Belarus; Clinical HIV Research Unit (CHRU), Wits Health Consortium (WHC), Department of Internal Medicine, School of Clinical medicine, Faculty of Health Sciences, University of Witwatersrand, Johannesburg, South Africa; Médecins sans Frontieres, Manson Unit, Public Health Department, London, United Kingdom; University of Liverpool, Department of Clinical Infection, Microbiology and Immunology, Liverpool, United Kingdom; London School of Hygiene and Tropical Medicine, Clinical Research Department, London, United Kingdom; Institute for Global Health, University College London, London, United Kingdom

**Keywords:** population pharmacokinetics, linezolid, rifampicin resistant tuberculosis, probability target attainment

## Abstract

Linezolid is a key component of the BPaLM regimen (bedaquiline, pretomanid, linezolid, and moxifloxacin), the recommended treatment for rifampicin-resistant tuberculosis (RR-TB). However, its optimal dosing and duration remain uncertain. To support dose optimization and efficacy interpretation, we developed a population pharmacokinetic (PK) model of linezolid and evaluated exposure and probability of target attainment (PTA). Ninety-four RR-TB patients received linezolid 600 mg daily for 16 weeks, followed by 300 mg daily for 8 weeks. Plasma samples were collected at multiple time points across 24 weeks and analyzed using high-performance liquid chromatography-tandem mass spectrometry. PK modeling was performed using nlmixr2 in R. A one-compartment model with first-order absorption and elimination, fat-free mass allometric scaling best described the data. Typical clearance and volume were 6.66 L/h and 58.8 L, respectively. Median AUC₀₋₂₄ was 84.34 mg·h/L at 600 mg and 46.36 mg·h/L at 300 mg. Median trough concentrations were 0.71 mg/L and 0.39 mg/L, respectively. The PTA analysis showed that the 600 mg dose achieved the fAUC₀₋₂₄/MIC target of 119 only for isolates with MICs ≤0.25 mg/L, however our randomized controlled trial findings suggest that a step-down strategy from 600 mg to 300 mg daily maintains efficacy in RR-TB treatment despite the lower linezolid exposure. This highlights the need to revise current pharmacokinetic-pharmacodynamic (PK-PD) targets, which are based on linezolid monotherapy, to better reflect outcomes in combination therapy. These data support the use of BPaLM regimens with reduced toxicity risk and provide a foundation for future dose optimization efforts.

## Introduction

Tuberculosis remains the deadliest single infectious disease worldwide, responsible for the deaths of 1.2 million people globally each year, and drug resistance continues to threaten progress in controlling TB(1). The World Health Organization recommends the six-month all oral bedaquiline, pretomanid, linezolid, and moxifloxacin (BPaLM) regimen as the prioritised treatment for rifampicin resistance TB. Moreover, all the other seven recommended rifampicin resistant (RR-)TB regimens include linezolid(2).

Linezolid is an oxazolidinone class antimicrobial originally approved for the treatment of Gram-positive bacterial infections. It inhibits ribosomal protein synthesis by binding to the 23S RNA peptidyl transferase centre of the 50S subunit of the prokaryotic ribosome(3). It is highly bioavailable and rapidly absorbed as an oral tablet of 600mg, and has a half-life of approximately 4 hours. It is partially metabolized in the liver, with non-renal pathways accounting for 65% of total clearance, while about 30% of the dose is excreted unchanged in the urine(4).

Early bactericidal activity studies in tuberculosis patients showed that linezolid doses between 300 mg and 1,200 mg kill *Mycobacterium tuberculosis* (*Mtb*) in sputum(5, 6). A study using the *in-vitro* hollow fiber system of infection to evaluate linezolid’s ability to kill *Mtb* at *in-vivo* mimicking conditions confirmed similar early bactericidal activity as observed in humans(7).

Linezolid pharmacokinetic (PK) - pharmacodynamic (PD) efficacy endpoints have however not been derived using patient data, and instead rely on *in-vitro* hollow fiber system of infection experiments investigating linezolid mono-therapy(8). These endpoints have subsequently been used for probability of target attainment (PTA) simulations in combination with linezolid population PK models that were developed on data from patients receiving combination therapy. It was concluded that a ≥90% PTA with 600 mg daily doses could only be achieved if the MIC of the *Mtb* was below 0.5-1.0 mg/l (9–12).

The optimal dose and duration of linezolid for treating tuberculosis in combination with bedaquiline, pretomanid, and moxifloxacin has consequently not been established through data-driven approaches. Especially explanatory PK-PD evidence to support an optimal linezolid dose has, until now, been lacking(13). In the meantime, clinical evidence increasingly supports a regimen of 600 mg daily for the first 16 weeks followed by 300 mg daily for the remaining 8 weeks of treatment, whilst 1,200 mg daily is associated with significant toxicity (2, 9, 14–16). However, the World Health Organization currently recommends 600mg for the full duration of treatment.

This study aimed to investigate linezolid PK properties amongst rifampicin resistant tuberculosis patients in the TB-PRACTECAL trial receiving combination therapy and to assess previously reported PK-PD endpoints against prospectively collected *Mtb* susceptibility data(17).

## Results

A total of 91 participants from Belarus and South Africa were included in the study. The median age was 36 years (range: 19 – 71 years) and 33 (36%) of the participants were female as shown in Table 1. Participants contributed a total of 952 timed plasma measurements, 14% (90/655) of the plasma samples had linezolid concentrations below the limit of quantification, and 655 of the plasma samples were measured between the first and last dose of treatment. Observed linezolid concentration ranged from 47ng/ml to 30,650 ng/ml with a median trough concentration of 20 ng/ml.

**Table 1:** Baseline characteristics of study participants.

|  |  |
| --- | --- |
| Sample size (n) | 91 |
| Age (y) | 36 (19-71) |
| Baseline weight (kg) | 56.6 (39.2-144.4) |
| Body mass index (kg/m<sup>2</sup>) | 19.8 (14.3-47.1) |
| Fat free mass (kg) | 45.2 (28.6-75.5) |
| BUN (mmol/L) | 3.6 (1.7-8.5) |
| ALT (IU/L) | 19 (4-113) |
| AST (IU/L) | 22 (4-82) |
| Total protein (g/L) | 77 (61-118) |
| Estimated creatinine clearance (mL/min) | 105.8 (43.4-243.8) |
| Female, n (%) | 33 (36.3) |
| Caucasian race, n (%) | 39 (42.9) |
| Black race, n (%) | 50 (54.9) |
| Asian race, n (%) | 1 (1.1) |
| Other race, n (%) | 1 (1.1) |
| HIV positive, n (%) | 36 (39.6) |
| BPaLM, n (%) | 38 (41.8) |
| BPaLC, n (%) | 29 (31.9) |
| BPaL, n (%) | 24 (26.4) |

Median (min-max) if not stated otherwise. BPaLM =
bedaquiline+pretomanid+linezolid+moxifloxacin, BPaLC =
bedaquiline+pretomanid+linezolid+clofazimine, BPaL =
bedaquiline+pretomanid+linezolid, ALT= alanine transaminase, AST= aspartate
aminotransferase. BUN = Blood Urea Nitrogen

A one compartment distribution model with first-order absorption and elimination best captured linezolid’s absorption, distribution and metabolism/elimination characteristics. Typical clearance and volume of distribution estimates were 6.66 L/hr and 58.8 L, whilst absorption rate constant was fixed to 1.23 hr^-1^ based on literature values(18). A lognormal model was used to estimate residual variability (Table 2 and Appendix 1).

**Table 2:**
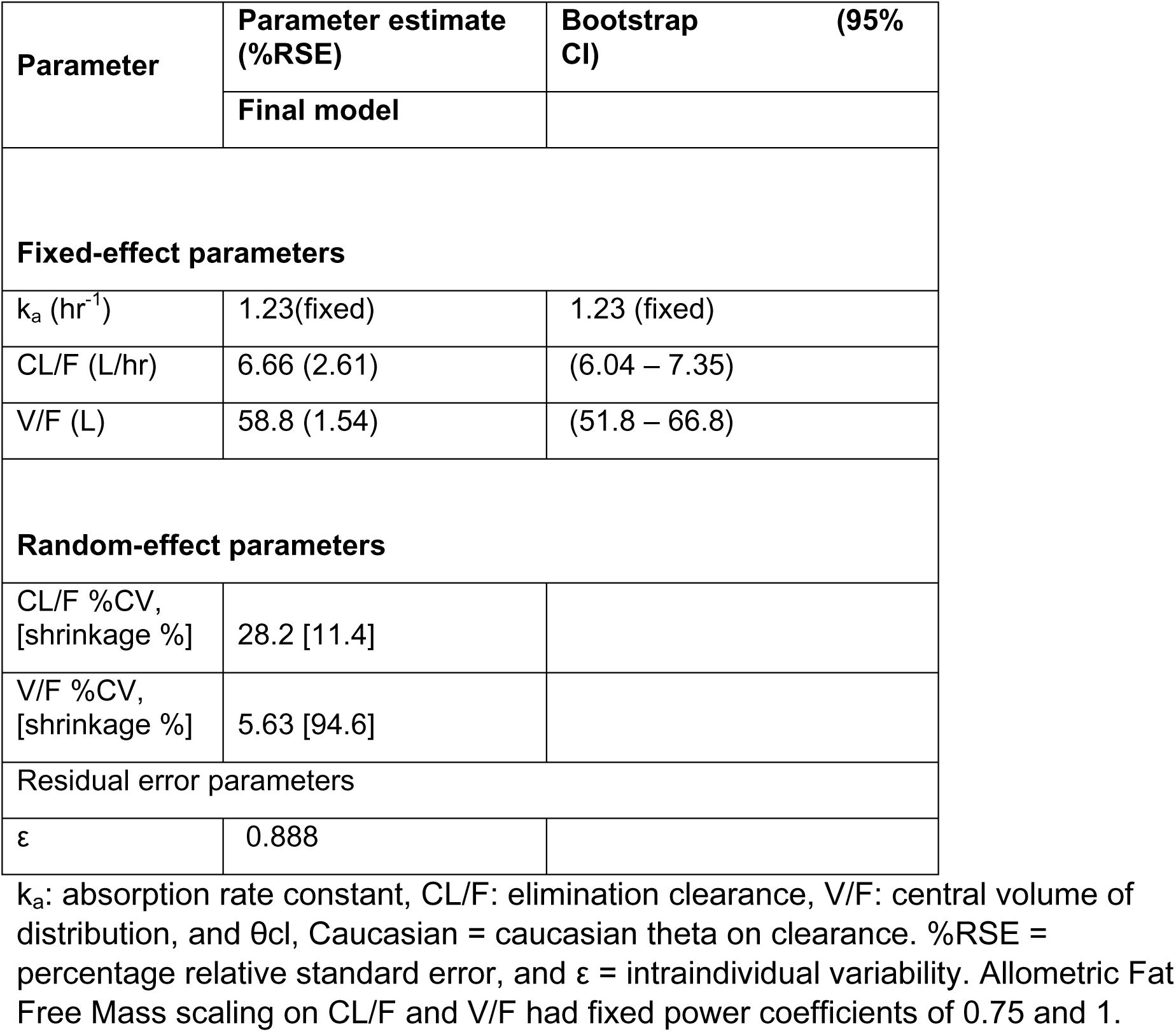
Estimated linezolid population pharmacokinetic.

Allometric fat free mass scaling for body size was embedded in the model a priori, and no further statistically significant covariates were identified using stepwise covariate modelling. Age, Caucasian race, Black race, and creatinine clearance on elimination clearance and black race and Caucasian race on volume were identified in the forward-step but could not be retained in the back-wards step (Appendix 2).

Goodness-of-fit (GOF) plots for the final model showed no significant bias from the unity line, indicating that the model predicted individual and population values closely matched the observed PK data (Figure 1). The time after first and last dose Visual Predictive Check (VPC) demonstrated satisfactory predictive accuracy of the developed model (Figure 2). Individual drug profiles were included in Appendix 3.

**Figure 1:**
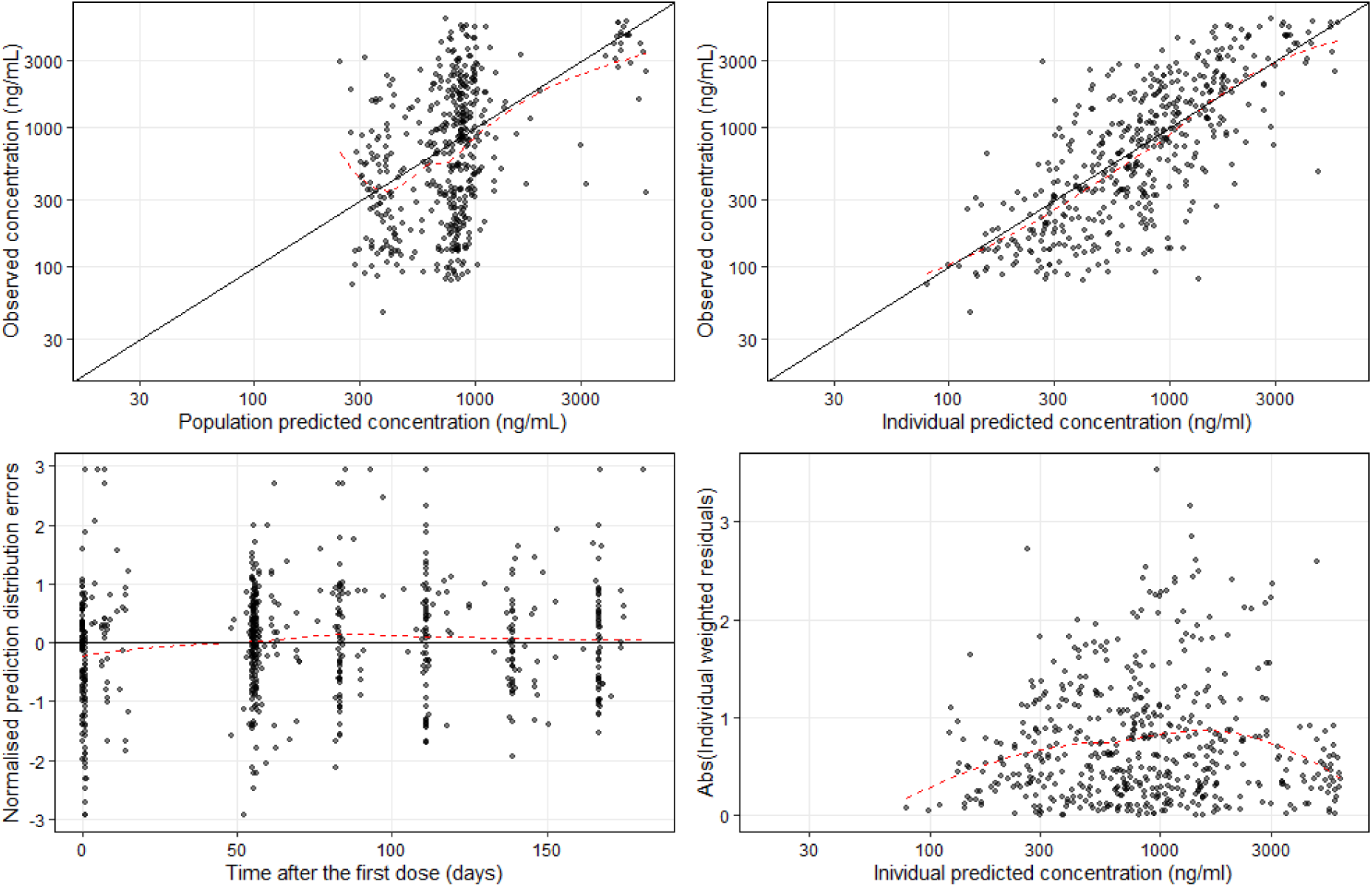
Linezolid population pharmacokinetic model goodness of fit plots. Dots represent observations, solid black lines represent identity lines and the red dashed lines represent locally estimated scatterplot smoothing.

**Figure 2:**
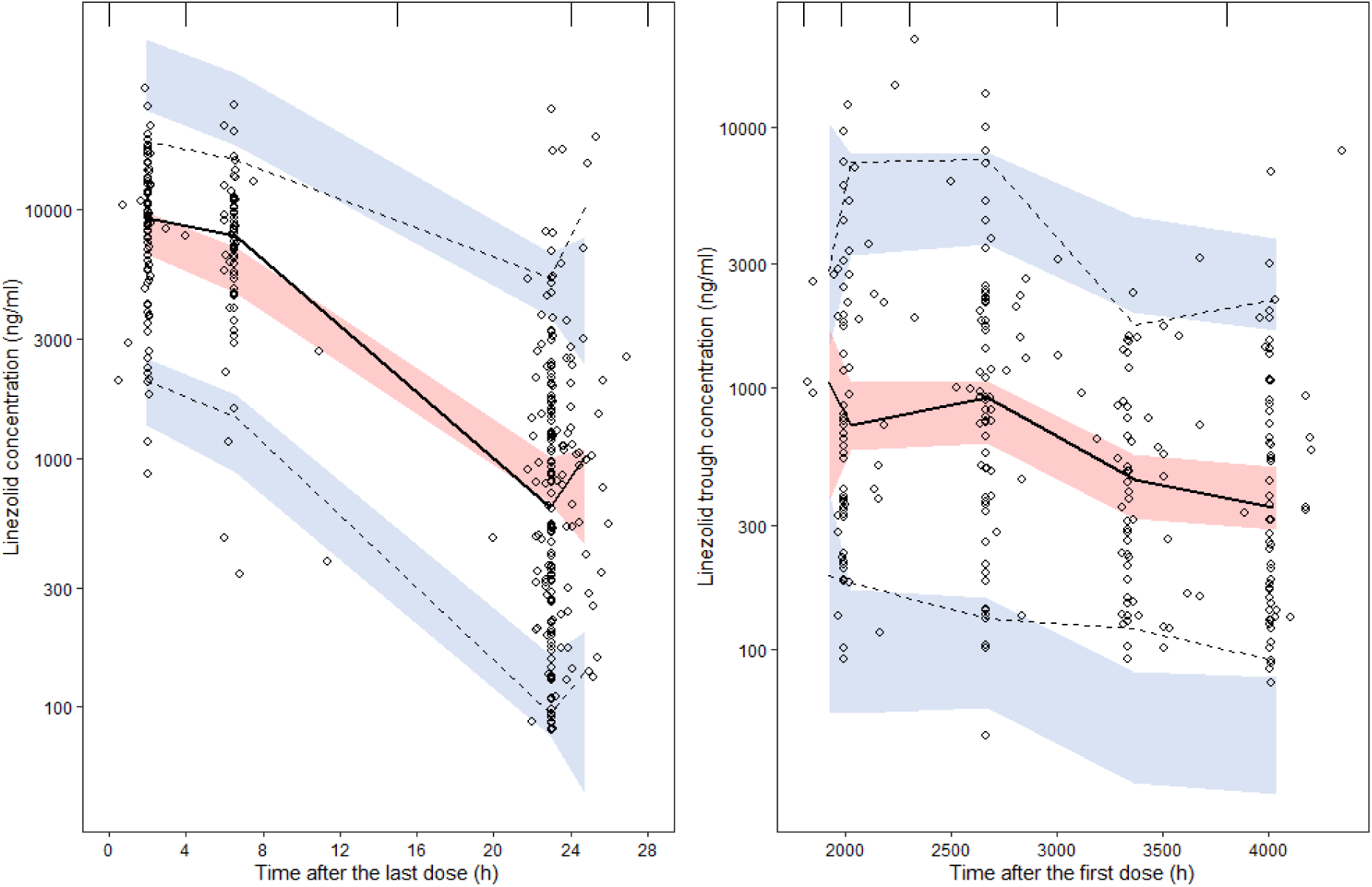
Visual predictive check, presented as time after last dose of the day 1 and week 8 visit (left panel) and time after first dose for the subsequent visits (right panel). Open circles represent the observed data, with dashed and solid lines presenting the 95% percentiles and median of the observed data. The blue and red shaded areas represent the 90% confidence intervals of model predicted 95% percentiles and median of the simulated data.

Robustness of model parameter estimates was verified using 1,000 non-parametric bootstraps. Median values of estimated parameters obtained using bootstrap analysis were consistent with corresponding final model parameter estimates, thus reflecting the final model’s robustness (Table 2). Linezolid population PK model derived secondary parameters maximum concentration (C_max_), area under the plasma concentration-time curve (AUC), and trough concentration (C_trough_) were presented in Table 3.

**Table 3:** Secondary pharmacokinetic parameters derived with the linezolid model.

| Parameter (unit) | Median | range |
| --- | --- | --- |
| **Day 1 600mg dose** |  |  |
| AUC 0-24 (mg hr/L) | 84.34 | 35.67-150.12 |
| Ctrough (mg/L) | 0.71 | 0.12 - 2.49 |
| Cmax (mg/L) | 8.03 | 4.48 - 12.79 |
| **Steady state 600mg dose** |  |  |
| AUC 0-24 (mg hr/L) | 85.76 | 35.68 - 156.88 |
| Ctrough (mg/L) | 0.71 | 0.12 - 2.98 |
| Cmax (mg/L) | 8.28 | 4.48 - 12.96 |
| **Steady state 300mg dose** |  |  |
| AUC 0-24 (mg hr/L) | 46.36 | 18.22 - 88.51 |
| Ctrough (mg/L) | 0.39 | 0.06 - 1.54 |
| Cmax (mg/L) | 4.48 | 2.29 - 7.04 |

Cmax: maximum linezolid concentration, tmax: time to reaching maximum linezolid concentration, ctrough: concentration just before the next dose was administered, and AUC: area under the plasma concentration-time curve.

Pure isolates of *M. tuberculosis* from 457 TB-PRACTECAL study participants had a median and mode MGIT MIC of 0.5mg/L with a range of <0.063 to 1mg/L as shown in Figure 3.

**Figure 3:**
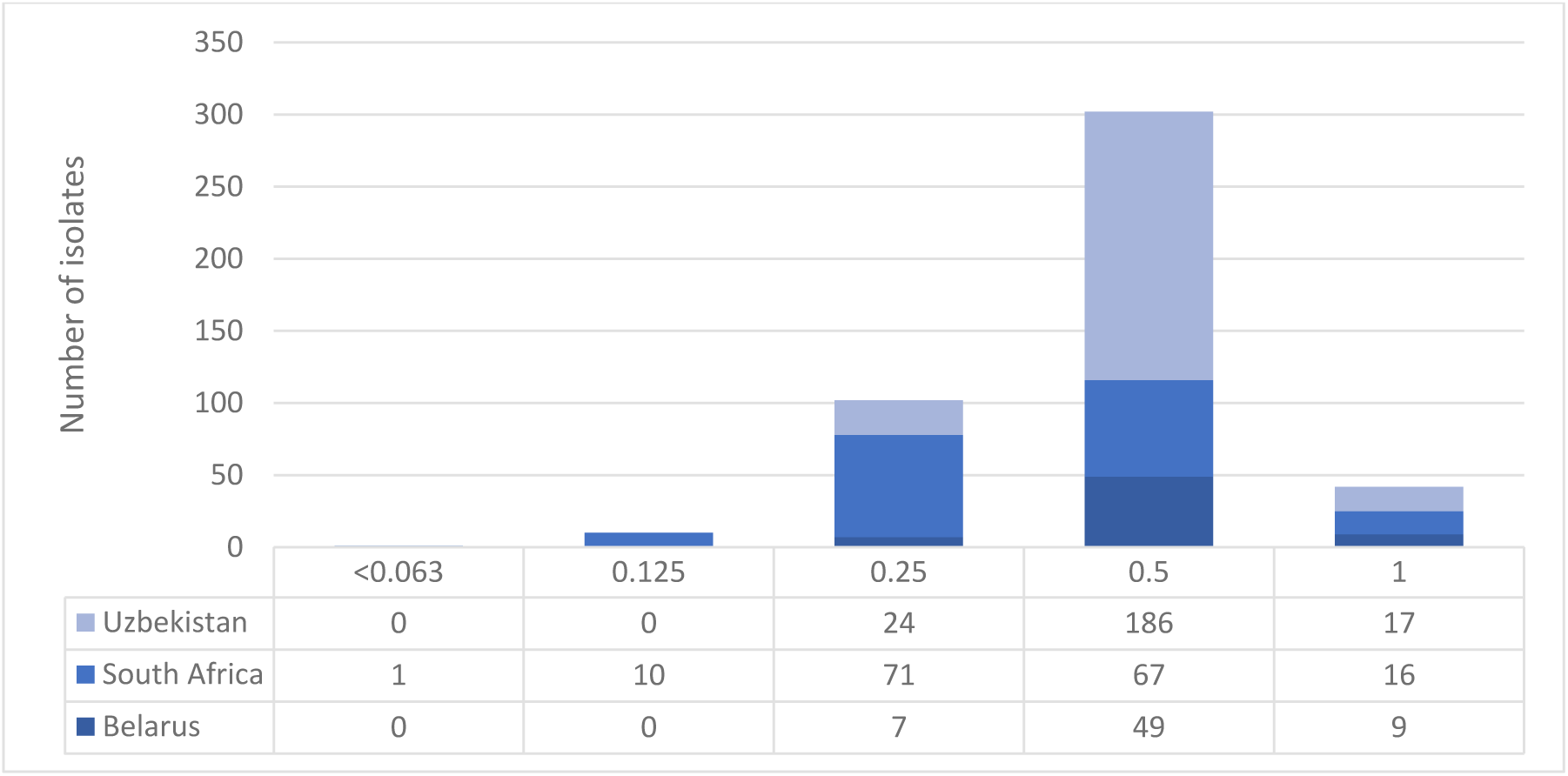
Distribution of pre-treatment isolates across linezolid MICs in the TB-PRACTECAL trial disaggregated by country of recruitment.

Using 2,000 stochastic simulations based on the observed individual characteristics of 94 patients, we evaluated the PTA for several PK-PD targets. These included the endpoint associated with 80% of maximal bacterial kill (EC80; fAUC₀₋₂₄/MIC ratio of 119) at day 28, the endpoint for a 1.0 log₁₀ bacterial kill (fAUC₀₋₂₄/MIC ratio of 73.60), the endpoint for sterilization at day 42 (fAUC₀₋₂₄/MIC ratio of 43.47), and the endpoint for bacteriostasis (fAUC₀₋₂₄/MIC ratio of 16.24)(8). Among these, only the bacteriostatic target (fAUC₀₋₂₄/MIC = 16.24) was achieved in ≥90% of simulations across all susceptible isolates with MICs up to 1 mg/L, for both the 300 mg and 600 mg linezolid doses. In contrast, the targets associated with 1.0 log₁₀ kill and sterilization at day 42 were only attained in ≥90% of simulations for isolates with MICs of 0.5 mg/L or lower, for both dosing regimens. Finally, the EC80 target at day 28 was only achieved in ≥90% of simulations for isolates with MICs of 0.125 or 0.25 mg/L, again for both the 300 mg and 600 mg doses (Figure 4).

**Figure 4:**
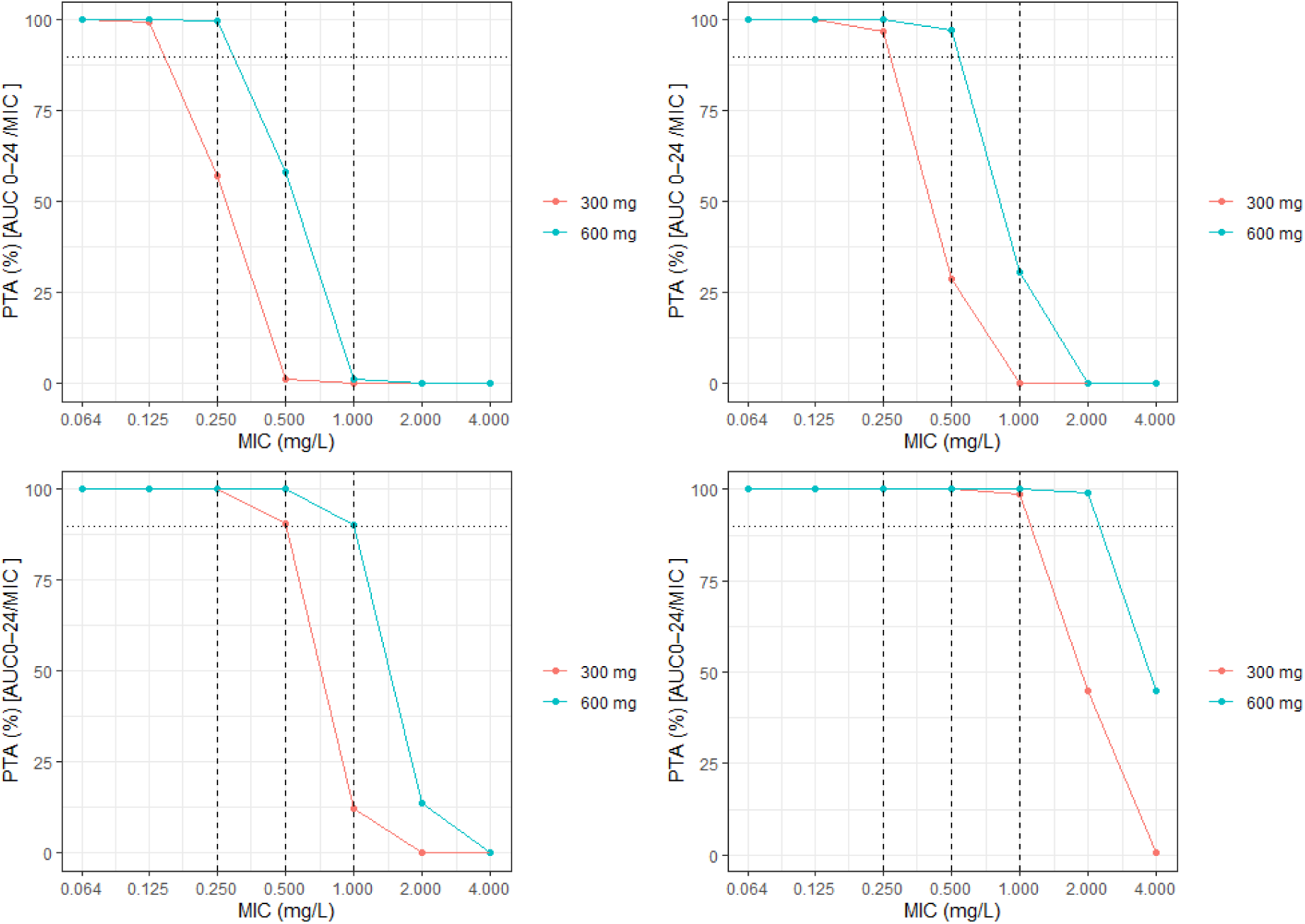
Plot of the day 14 probability of fAUC/MIC target attainment of 119 for 80% of maximal bacterial kill at day (top left), 73.6 for a 1.0 log10 bacterial kill (top right) 43.47 for sterilization at day 42 (bottom left) and 16.24 for bacteriostasis (bottom right). Red and blue lines represent 300 and 600 mg linezolid dosing, respectively.

## Discussion

A one-compartment disposition model with first order absorption and elimination best described linezolid population PK in rifampicin resistant tuberculosis patients from South Africa and Belarus being treated with BPaL-based short regimens. Varying approaches have been reported for characterizing the absorption phase, including transit compartments or lag functions(19, 20). There was only one 2-compartment model and one accounted for autoinhibition of elimination(12, 19).

Linezolid clearance in TB patients has been reported to range between 3.81 and 7.69 L/hr and our estimate at 6.66 l/hr falls on the higher end of this range(4, 9, 11).

Consequently, our median AUC_0-24_ for the 300mg and 600mg daily doses at 46.36 mg*hr/L and 85.76 mg*hr/L, respectively, tend to be at the lower end compared to previously reported values (Table 3)(4, 9, 21).

Previously published covariates influencing linezolid population PK parameters are body size-related such as fat free mass, which we included in our model, and weight(12, 18, 19). Variation in kidney functioning such as creatinine clearance and blood urea nitrogen have either been identified in model development or have been included in allometric adjustment(10, 12, 18). Creatinine clearance was identified in the first cycle of the stepwise covariate search in our model at *p<0.05*, although it was dropped in during the backward elimination step at *p<0.001*. This may have been caused by the strict inclusion criteria for our trial, where patients with severely impaired renal function were excluded. Diabetes mellitus type 2 has also been reported as a covariate, but this could have been explained by its effect on renal functioning and was not found to be a significant confounder in our model(12).

Consistent with what has been reported elsewhere, HIV infection did not significantly influence our model(9). We identified race as influencing both volume and clearance in the forward step of the covariate model building. This may be explained by genetic variability which was also reported in our study population; Variant *3 in CYP3A5*3 was associated with lower linezolid trough values(22). Linezolid volume of distribution has been reported to range from the 31 L to 50 L, and we reported a volume close to the higher end(19, 20, 23).

The linezolid median MIC in MGIT in the study population was 0.5 mg/L. This is similar to the distribution reported by Abdelwahab et al. but at almost one dilution step higher than Zhang et al.(9, 12). Half of the isolates from South Africa had an MIC equal to or below 0.25mg/L while for Belarus and Uzbekistan this was 11%. As this difference is just one dilution apart, and despite stringent standards implemented in the trial, these could be analytical differences as three different laboratories were used.

Target attainment for EC80 target at day 14 in our study was only achieved in ≥90% of simulations for isolates with MICs of 0.125 or 0.25 mg/L, for the 300 mg and 600 mg doses. This was within the range of previously reported breakpoints at 0.125-0.5 and 0.25-1.0 mg/L in different studies (9, 11, 12).

The median Ctrough of 0.71 mg/L for the 600mg dose and 0.39 mg/L for the 300mg dose in our study was below 2 mg/l (Table 3). The low Ctrough may explain the very low linezolid related adverse events reported in the TB-PRACTECAL trial as a threshold of 2mg/L has been defined as associated with low toxicity(9, 17, 18).

Linezolid dose optimization data is sparse when investigated in the context of drug combinations such as BPaLM or BPaLC, where the companion drugs may play different roles due to their variability in penetration of the different TB lesions(23). Our results show, in line with previous reports, that PTA simulations tend to underestimate the efficacy of linezolid when given in combination with potent companion drugs. Whilst simulations often report undesirable attainment for higher MICs, our study showed linezolid-based combination therapy to be highly effective in patients. This emphasizes that the current PK-PD targets, that have thus far been derived using *in-vitro* hollow fiber infection experiments using linezolid monotherapy, do not extrapolate to *in-vivo* settings where linezolid is administered with companion drugs such as bedaquiline, pretomanid, and moxifloxacin or clofazimine.

The main limitation of the study is related to the fact that it was a sub-study, and thus the sample size was opportunistic with the sampling schedule being selected using an optimal design approach(24). Moreover, the estimated total sample size was not achieved, due to early discontinuation of the main study as a result of reaching non-inferiority to the standard of care(15). Due to the absence of data driven linezolid efficacy endpoints, PTA simulations lack translational power for informing drug combination therapy in the clinic.

As BPaLM, BPaLC and BPaL regimens have been shown to be highly efficacious(17), our study provides explanatory evidence that even relatively low linezolid exposure may have contributed to these outcomes. These results reignite the need to explore combined PK-PD efficacy and safety targets for linezolid and other new oxazolidinones when used as part of efficacious regimens.

## Materials and Methods

This was a sub study nested in the TB-PRACTECAL randomised controlled trial in patients with rifampicin resistant tuberculosis (ClinicalTrials.gov NCT02589782). Participants in Belarus and South Africa receiving one of three investigational regimens could participate in this sub study. The BPaL arm consisted of bedaquiline 400mg daily for 2 weeks then 200mg three times a week for 22 weeks, pretomanid 200mg daily for 24 weeks and tapered dose linezolid 600mg daily for 16 weeks then 300mg for 8 weeks. Clofazimine 100mg daily for 24 weeks was added in BPaLC arm or Moxifloxacin 400mg daily for 24 weeks in BPaLM arm. Blood samples were collected on Day 1 (0, 2 and 23 hours), Weeks 8 (predose, 6.5 and 23 hours), and predose at the 12, 16, 20, 24, 32 and 72 week post randomisation visits. Drug concentrations were quantified in a GCP laboratory using a high-performance liquid chromatography-tandem mass spectrometry. The lower limit of quantification for linezolid was 80ng/mL.

Linezolid population PK data was analyzed using a non-linear mixed effect modelling approach. Nlmixr2, an open-source R package was used for population PK modelling and simulation estimation. R v4.4.1 was used for dataset creation, data exploration and generation of tables and plots (Appendix 4). The first-order conditional estimation with interaction (FOCE-I) algorithm in nlmixr2 was used. Inter-individual variability (IIV) at the parameter level and residual variability (RV) at the observation level was used to dissect and quantify variability between patients from residual variability.

In line with previous reports in literature one and two-compartment linear models were evaluated with combined, proportional and additive residual error models or a log-transformed residual error model. Random effects on clearance (CL) and volume of distribution (V) were included in the model. Various absorption models were explored including transit compartment models and fixed absorption constant (ka).

A covariate matrix, including age, sex, weight, body mass index, fat-free mass, race, BUN, ALT, AST, total protein, creatinine clearance, treatment regimen, and eta estimates for clearance and volume of distribution from the base model, was used to explore both correlations and potential collinearity among covariates. Fat free mass allometric scaling was applied to both volume of distribution and clearance. The coefficients of the power model were fixed to 1s for V and 0.75 for CL. The selected covariates underwent stepwise forward inclusion (P<0.05, ΔOFV > 3.84 per degree of freedom) and backward elimination (p<0.001, ΔOFV> 10.83 per degree of freedom) to select those that would improve the model fit significantly.

Goodness-of-fit plots were used to assess how well the model predicted individual and population values closely matched the observed PK data. Model validation was also performed using visual predictive check (VPC, n=2,000) plots and bootstrapping (n=500)(25, 26). The shrinkage, relative standard error, and variability value including omega and sigma values were also used to assess the precision and robustness of the model.

Minimum inhibitory concentrations were determined from a routine testing concentration set (1, 0.5, 0.25 mg/L) in Mycobacterial Growth Indicator Tubes; testing was performed using a higher (32, 16, 8, 4, 2 mg/L) or lower (0.125, 0.016 mg/L) testing concentration set if required. The results from all participants from the TB-PRACTECAL trial were summarised by country of enrolment and the median and interquartile range were reported.

PTA for the various efficacy targets was explored at daily oral intake of 600 mg linezolid in first 16 weeks followed by 300 mg in the latter 8 weeks. The PK-PD breakpoint was defined as the highest MIC at which the PTA is >90%. Stochastic simulations (n = 2,000), with patient level characteristics from the observed participants in this study, accounted for protein binding at 31%(27). Selected endpoints were fAUC₀₋₂₄/MIC ratio of 119 for 80% of maximal bacterial kill (EC80) at day 28, fAUC₀₋₂₄/MIC ratio of 73.60 for a 1.0 log₁₀ bacterial kill, fAUC₀₋₂₄/MIC ratio of 43.47 for sterilization at day 42, and fAUC₀₋₂₄/MIC ratio of 16.24 for bacteriostasis(8).

## Data Availability

All data produced in the present study are available upon reasonable request to the authors

## Acknowledgements

FK conducted the research as part of a Sir Henry Dale Fellowship jointly funded by the Wellcome Trust and the Royal Society (Grant Number 220587/Z/20/Z).

## Contributors

Conception and design: B-TN, FK, GD and DAM. Data acquisition: IM, RM, VS, SR. PK modelling: BT-N and FK. First draft of manuscript: BT-N. Revising and approval of manuscript: all

## Ethics statement

The study was approved by the MSF Ethics Review Board (reference no. 1541) and the LSHTM Ethics Committee (reference no. 16249). The Belarus RSPCPT ethics committee and the regulator-Centre of Excellence for the Minsk site. PharmaEthics for the Don Mckenzie and Dorris Goodwin hospitals sites, University of Witwatersrand Human Research ethics committee for the Helen Joseph and King DiniZulu Hospitals sites and the South Africa Health Products Regulatory Authority

## Data access statement

Deidentified data will be available to researchers upon a written request to the Medical Director, Médecins sans Frontières, Operational Centre Amsterdam, the Netherlands.

## Appendix 1: R code for the linezolid model

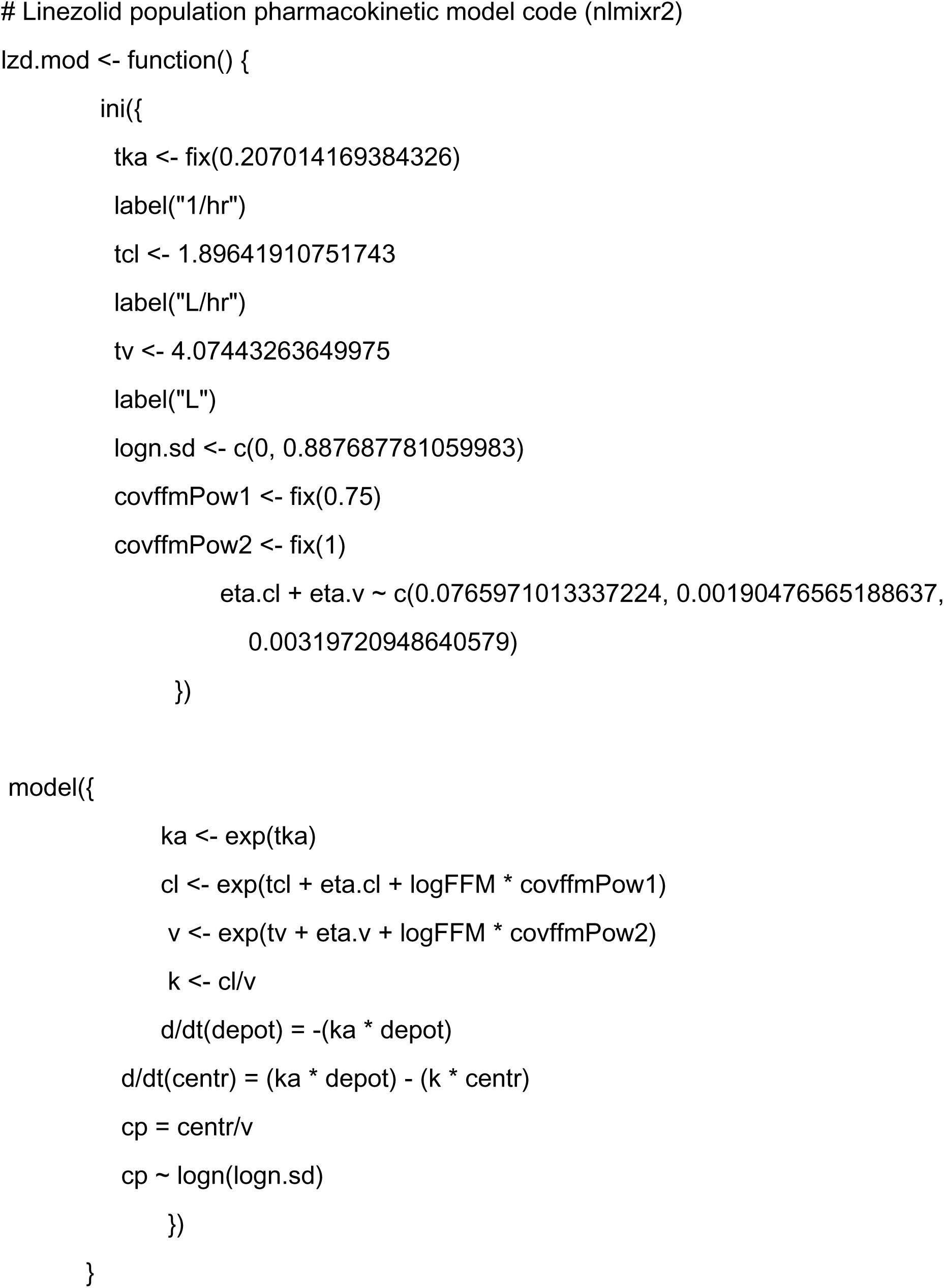

## Appendix 2: Covariate and parameter correlation matrix

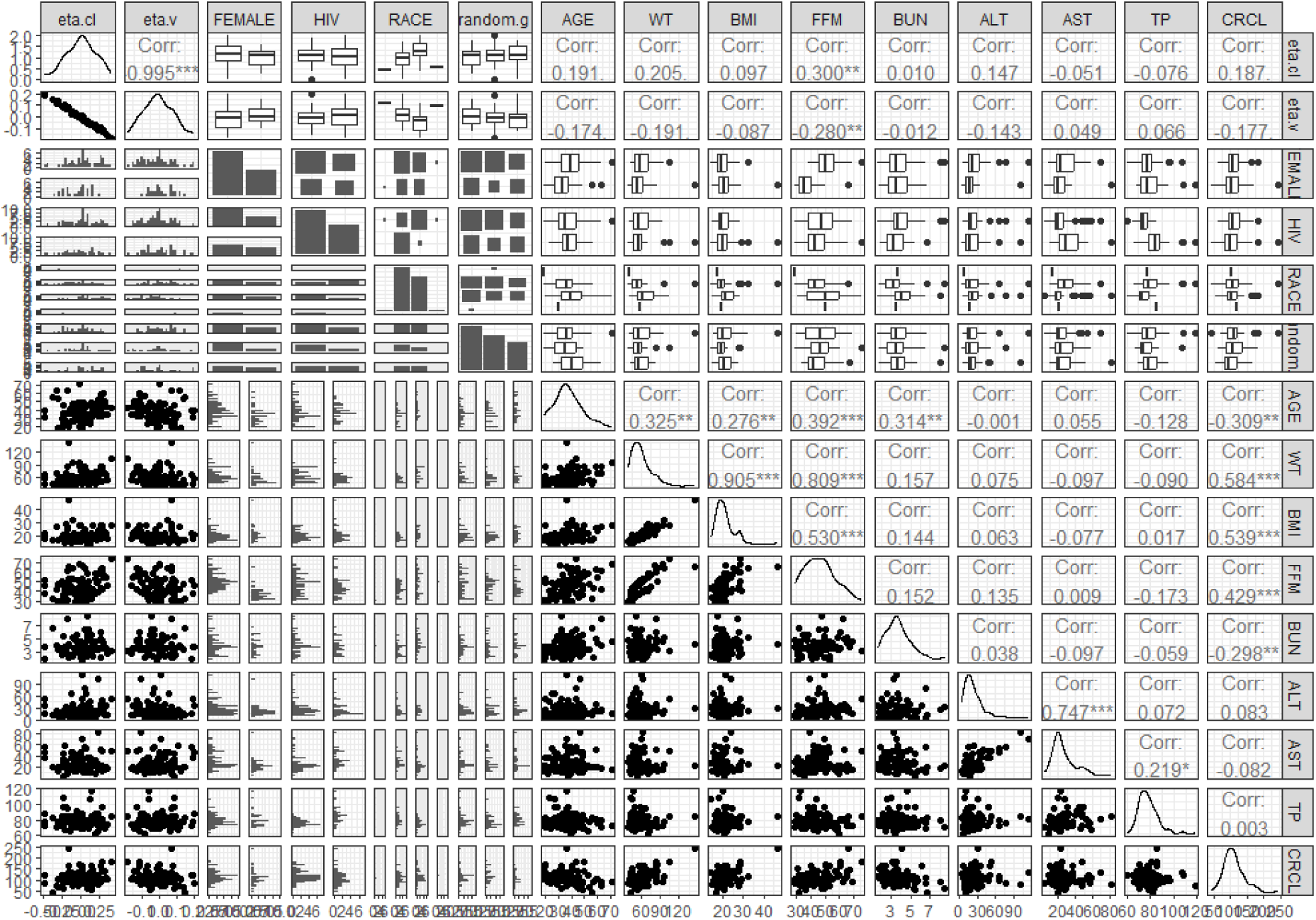

Covariate vs parameter matrix on base model. WT: weight, BMI: body mass index, FFM: fat free mass, BUN: blood urea nitrogen, ALT: alanine transaminase, and AST: aspartate aminotransferase, TP: total protein, CRCL: estimated creatinine clearance, random.g: BPaLM, BPaLC or BPaL arm.

## Appendix 3: Individual model fit plots

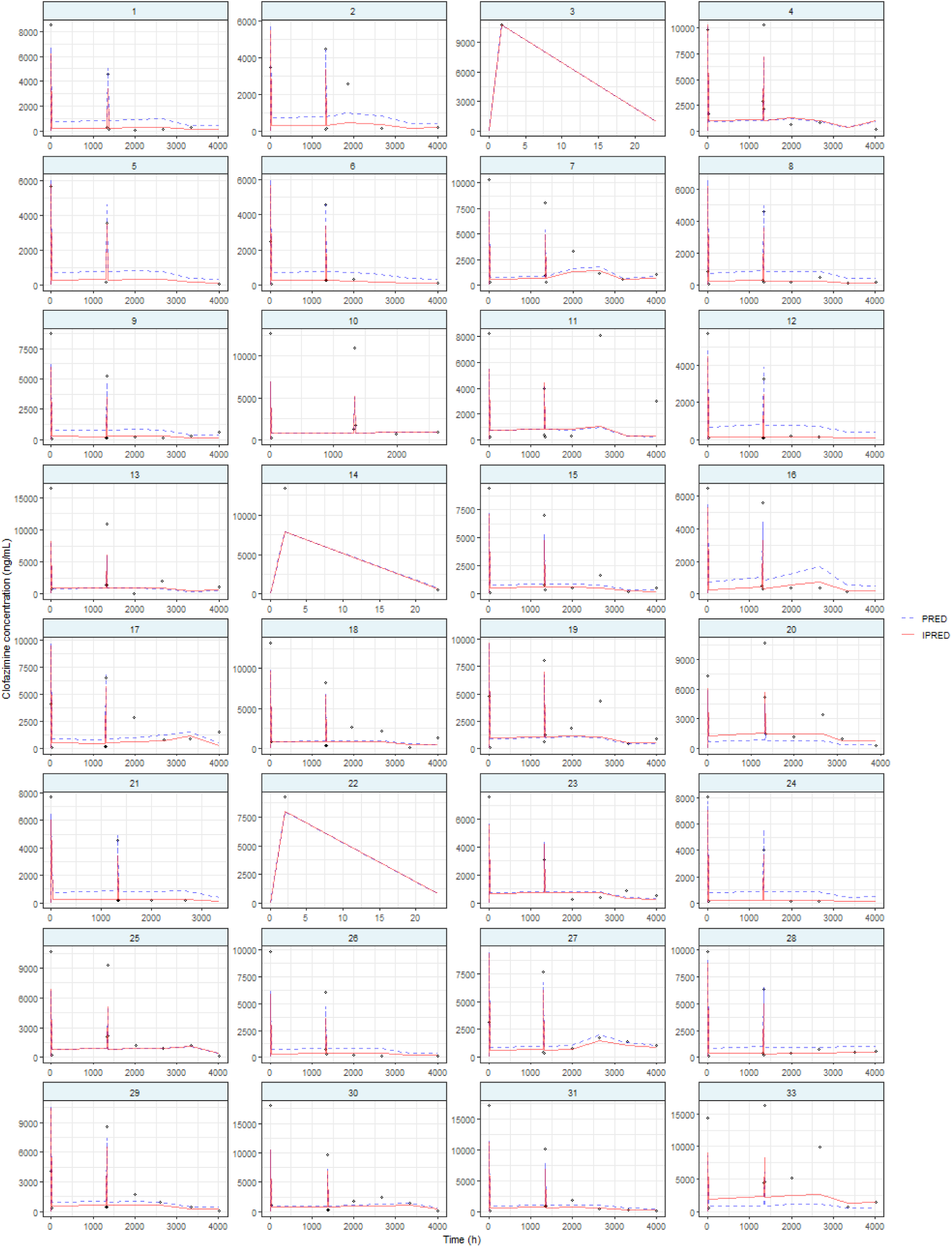
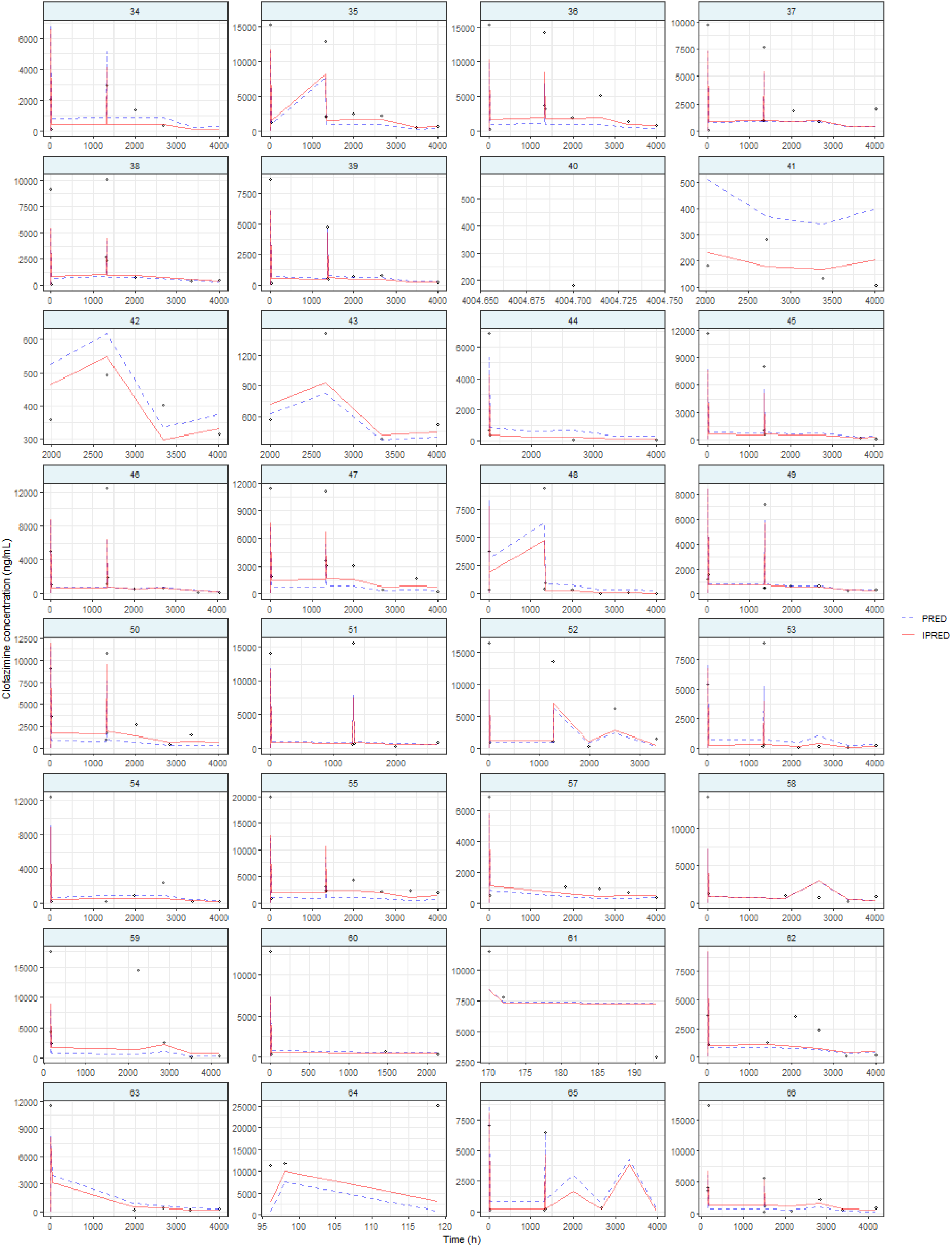
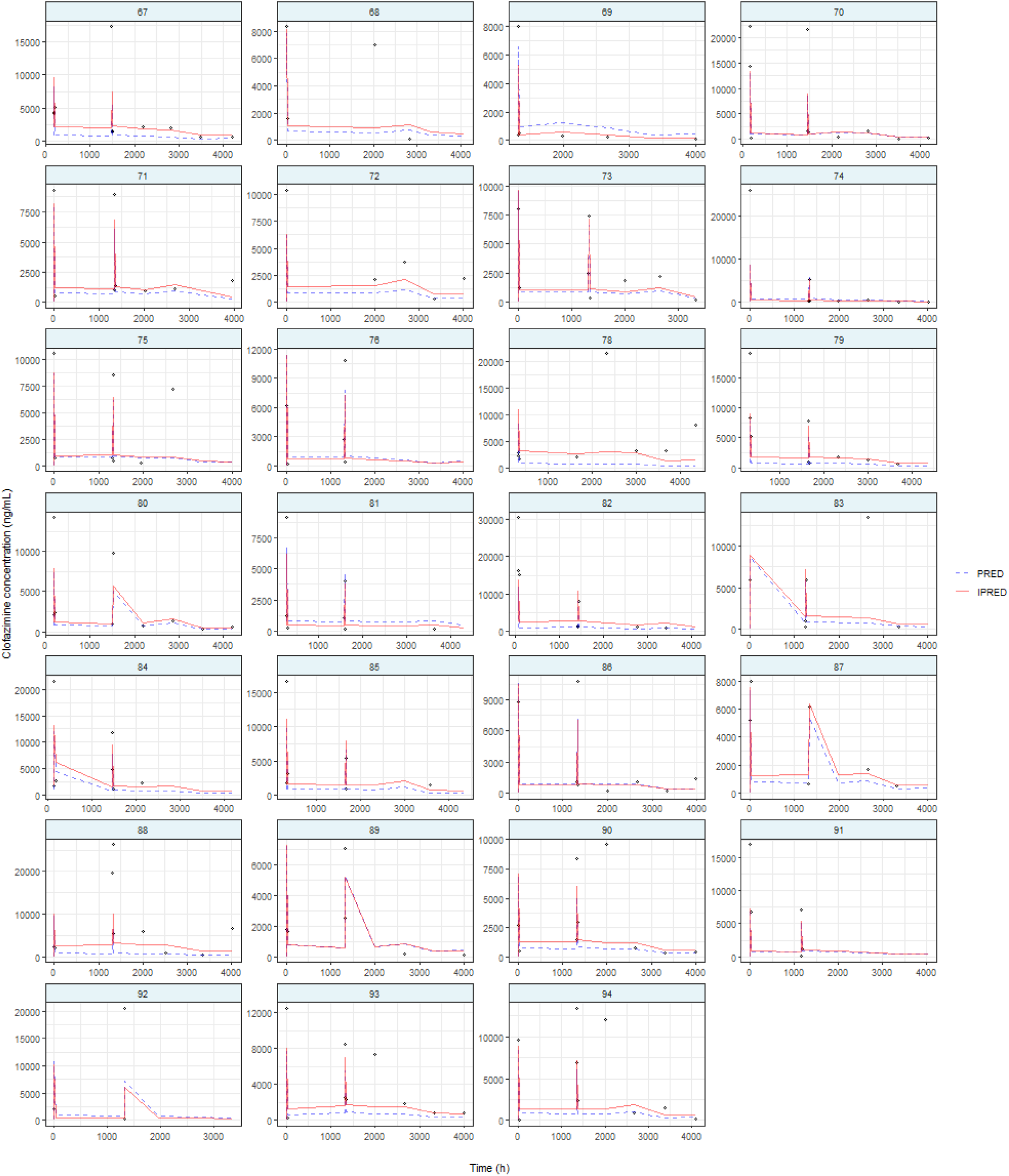

Individual linezolid plasma concentration - time profiles. Each panel represents a participant, with open circles representing observed plasma concentrations, the blue dashed line the population predictions by the developed model and the red solid lines the individual population predictions.

## Appendix 4: Used r packages

library(rxode2)

library(nlmixr2)

library(reshape2)

library(ggplot2)

library(tidyverse)

library(PerformanceAnalytics)

library(psych)

library(dplyr)

library(GGally)

